# Impact of COVID -19 on Depression and Anxiety among Healthcare Professionals in Abu Dhabi

**DOI:** 10.1101/2021.04.11.21253563

**Authors:** Amal Abdul Rahim Al Zarooni, Aljazia Khalfan Alghfeli, Hamda Musabbah Alremeithi, Roqayah Abdulla Almadhaani, Latifa Baynouna Alketbi

## Abstract

COVID-19 have affected Healthcare workers is many ways. One of the important areas is the psychological impact. The aim of this study is to examine the effects of the COVID-19 outbreak on the mental health of healthcare Professionals (HCP). A cross-sectional study was conducted between April 11^th^, and July 23^rd^, 2020, to assess depression and anxiety of healthcare workers, during the COVID-19 pandemic. An online, self-administered, anonymous questionnaire evaluated 1,268 HCP. More than half of the participants reported symptoms of anxiety (51.5%). Mild anxiety was reported in 28.8% of participating HCP, and 12.68 % of the participants registered moderate anxiety scores, while 9.95 % reported severe anxiety. Depression symptoms were revealed in 38.3 % of participating providers. Among all participates, 4.3 % and 2.7 % reported moderately severe and severe depression, accordingly, while 22.5%, and 8.8 % of the participating health care providers documented mild and moderate depression. The high prevalence of anxiety and depression recorded among HCP during the pandemic suggests that mental health intervention and support are necessary to ensure the psychological well-being of HCP.

## Introduction

The Ongoing infections of COVID-19 have created a global health burden and public health problems in many countries.^1^ COVID-19 was announced as a global pandemic on March 11, 2020 by the World Health Organization (WHO). COVID-19 is a highly contagious virus causes mild to severe respiratory tract infections.^2^ In the United Arab Emirates (UAE), changes in the health care system have been adapted to the public health needs caused by the COVID-19 pandemic. Currently, health care providers are making remarkable efforts to control additional outbreaks of the disease.^1^ Health care providers are the frontline during COVID-19 with higher risk of exposure to and acquiring COVID-19 infection, this may exposed them to mental distress causing anxiety, depression and emotional stress.

Providing direct care to patients with COVID-19, or being required to undergo quarantine or isolation, may lead to psychological distress in health care providers.^3^ Qualitative data in UAE frontline staff showed Covid-19 related stress, arising from working in hazard environment and the trauma from amount of death initially.^4^Infectious disease outbreaks are likely to affect the psychological health of healthcare workers (HCWs) practicing in the front lines of the pandemic.^5^Initial evidence that a considerable proportion of HCWs experienced mood and sleep disturbances during the outbreak, justifies the recommendation to establish methods to mitigate mental health risks and adjust interventions under the conditions of the pandemic.^6^Health care workers contributed significantly to controlling the infectious disease, in this context psychologist support, mental counselling, and helpline support were provided to the health care team in United Arab Emirates. The aim of the current study was to examine the effects of the COVID-19 outbreak on the manifestation of depression and anxiety in HCWs

## Methods

### Study design, setting, and population

A cross-sectional study was conducted in the United Arab Emirates, among health care providers from primary health care centers and inpatient hospitals in private and public sectors. The study was conducted between 11 April to 23 July. Due to the convenient online distribution of the survey, the heath care provider in Abu Dhabi Healthcare Servies (Seha) was determined, calculating a margin of error of 5%, and a confidence interval (CI) of 95%.

### Survey design

Data was collected using an anonymous online self-administered questionnaire, written in English. The survey was designed to collect information regarding demographic data of the participants, including age, sex, occupation, specialty, years of experience, the city and setting of their practice.

Depression screening was accomplished using the Nine-Item Patient Health Questionnaire (PHQ-9). The PHQ-9 includes nine items, each requiring four responses. Each response is scored according to the following: not at all=0, several days=1, more than half a day=2, almost every day=3. The total depression score may range from 0–27. A total score of 0–4 indicates minimal depression, with a score of 5–9 signifying mild depression. A total score of 10–14 indicates moderate depression, with scores of 15–19, and 20–27, demonstrating moderately severe and severe depression, respectively.

The Generalized Anxiety Disorder scoring system (GAD7) includes a 7-item questionnaire. Each item requires four responses, and each response is scored as follows: not at all=0, several days=1, more than half a day=2, almost every day=3. The total depression score may range from 0–21. A total score of 0–4 indicates minimal anxiety, with a total score of 5–9 demonstrating mild anxiety. Total scores of 10–14, and 15–21 indicate moderate and severe anxiety, respectively.

### Statistical analysis

Data were analyzed using SPSS version 21 Software program, Descriptive statistics such as mean, standard deviation (SD) were computed for quantitative variables and frequencies, and percentages were calculated for categorical variables. Linear and logistic regression analysis was conducted to determine the determinants. Significant level of p value ≤0.5 was used.

### Ethics and confidentiality

All study participants were completely informed about the purpose, methods, time frame, and their role in the study. Online consent for participation was established prior to enrolment.

The study was approved by the Ambulatory Health care services human ethics committee and the Abu Dhabi health care service central human Ethics Committee.

The study was conducted in accordance with the Ethics Committee’s guidelines. The questioner was anonymous to the participants and did not record any identifiers or personal information of the participants. Confidentiality of the study participants was maintained.

## Results

A total of 1,268 healthcare providers responded to the survey, 47.7% of whom (605) were working in in-patient hospitals, 19.6 % (249) in primary health care practices, and 15.9 % (202) were working in emergency and ICU care settings. This excellent response rate was achieved during the COVID-19 pandemic, between April 11^th^ and July 23^rd^, 2020. Majority of the participants (88.6 % [1,124]) were from SEHA, and only 144 originated from outside SEHA. Participants derived a mean age of 40.26 years and were mostly female (74.5%). Nurses constituted 61.9% of the respondents, and 18.4% were consulting physicians, specialists, and residents. Technicians comprised 14.7% and only 4.9% were pharmacists. Physicians who enrolled represented various specialties, including family medicine (6.2 %), internal medicine (7.9 %), Obstetrics and gynecology (8.3 %), Pediatrics (1.1 %), Psychiatry (1.7%), and Surgeons (9.5 %). Most participants originated from Abu Dhabi and Al Ain (62.5 %, 26.1 % respectively), and 9.9 % of participants hailed from the western region. The mean work experience was 14.3 years. (Table 1)

**Table 1.**
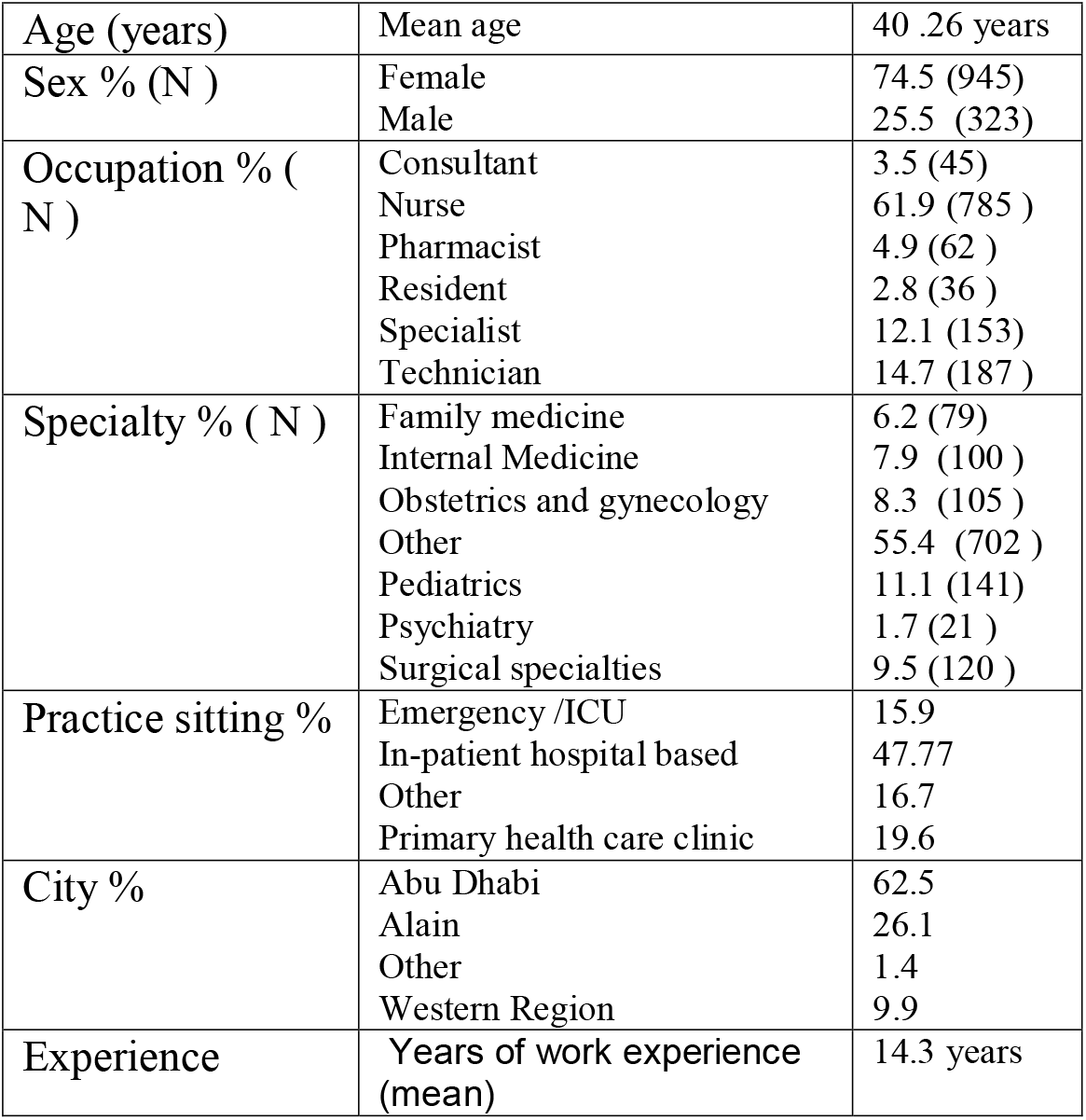
Characteristics of the study population.

| **Table 1. Characteristics of the study population.** |  |  |
| --- | --- | --- |
| Age (years) | Mean age | 40 .26 years |
| Sex % (N ) | Female | 74.5 (945) |
|  | Male | 25.5 (323) |
| Occupation % ( N ) | Consultant | 3.5 (45) |
|  | Nurse | 61.9 (785 ) |
|  | Pharmacist | 4.9 (62 ) |
|  | Resident | 2.8 (36 ) |
|  | Specialist | 12.1 (153) |
|  | Technician | 14.7 (187 ) |
| Specialty % ( N ) | Family medicine | 6.2 (79) |
|  | Internal Medicine | 7.9 (100 ) |
|  | Obstetrics and gynecology | 8.3 (105 ) |
|  | Other | 55.4 (702 ) |
|  | Pediatrics | 11.1 (141) |
|  | Psychiatry | 1.7 (21 ) |
|  | Surgical specialties | 9.5 (120 ) |
| Practice sitting % | Emergency /ICU | 15.9 |
|  | In-patient hospital based | 47.77 |
|  | Other | 16.7 |
|  | Primary health care clinic | 19.6 |
| City % | Abu Dhabi | 62.5 |
|  | Alain | 26.1 |
|  | Other | 1.4 |
|  | Western Region | 9.9 |
| Experience | Years of work experience (mean) | 14.3 years |

Anxiety scores were measured for each of the 1,268 participants of healthcare providers. Generalized anxiety scores were measured using Generalized Anxiety Disorder-7. More than half of the participants (51.5%), reported symptoms of anxiety, and almost half of them (48.48 %) scored in the minimal anxiety category. Mild anxiety was reported in 28.8% of all healthcare providers, and 12.68 % respondents revealed moderate anxiety scores, with 9.95 % reporting severe anxiety.

Mild and moderate anxiety (22.6%, 9.9%) were noted among female healthcare providers. Slightly higher severe anxiety (6.8%) was noted among male healthcare providers although this was not significant.

It was noted that anxiety scores decreased with additional years of experience. Only 3.6% of healthcare providers with more than 30 years of experience reported severe anxiety, while 16% of HCP with less than 5 years of experience had scores indicating severe anxiety.

Age groups were correlated with anxiety scores among healthcare providers. Scores indicating more severe anxiety among healthcare provider of less than 30 years of age (14.5%). Mild anxiety was primarily observed among healthcare provider between 41–50 years of age (31.3%), while moderate anxiety was noted among healthcare providers who were beyond 50 years of age (16.2%).

Anxiety scores were significantly associated with occupation, p = 0.02. Mild anxiety affected more than 30% of the Nursing (30.2%), specialist, (32.5%), and resident (36.1%) occupations. Moderate anxiety was mostly noted among consultants (15.6%), and severe anxiety was significantly noted among residents (19.4%).

Anxiety scores were significantly associated with specialties, and p value was 0.029. Mild anxiety affected more than 30% of the Psychiatry (30%) Family medicine (30.4%), Pediatrics (31.9%), Obstetrics and gynecology, (35.6%) and Internal medicine (36.4%) specialties. Moderate anxiety was mostly noted among Obstetrics and gynecology (18.3%). Severe anxiety scores were the highest among Internal medicine (16.2%) specialists (See Table 2).

**Table 2:**
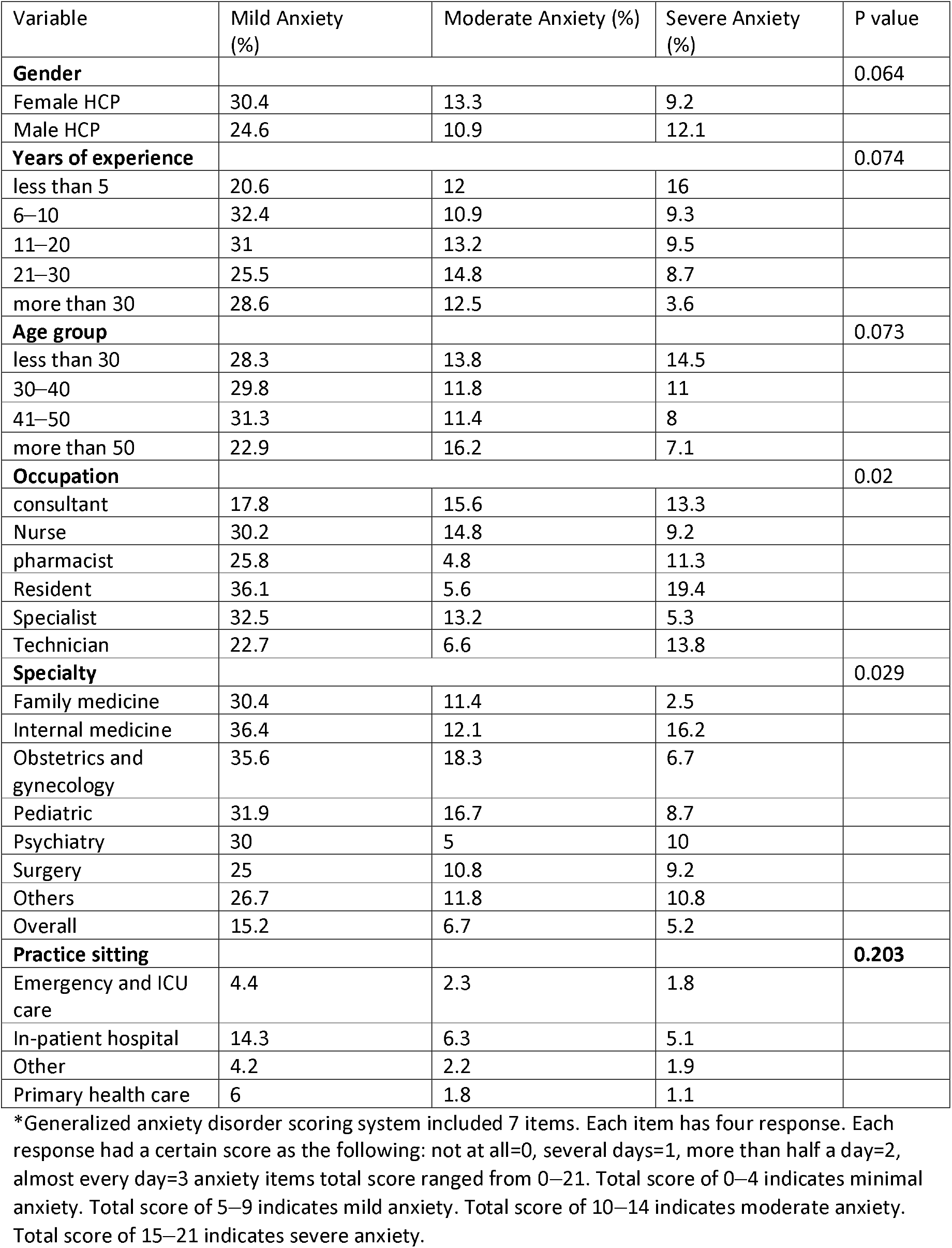
**Anxiety scores and prevalence among healthcare providers in correlation with HCP gender, years of experience, Age group, Occupation, and Specialty.**

Depression scores were measured for each of 1,268 participating healthcare providers. Depression symptoms were reported in 38.3% of respondents. Most of participants revealed minimal depression scores (60.3%). Moderately severe depression (4.3%) and severe depression (2.7%) were both less than 5%. HCPs reported 22.5% and 8.8 % in mild and moderate depression, respectively

Mild and moderate depression (23.5% and 9.1 %) respectively, were observed more frequently among females. Moderately severe (6.6%) and severe depression rates (3.5%) were higher among male healthcare providers. P value was not significant for depression severity in correlation with healthcare providers’ gender.

Mild, moderate, moderately severe, and severe depression were mostly noted among healthcare providers aged less than 30 (23.9%, 13.2%, 4.4% and 4.4%). High rates of moderately severe depression equivalence to that found among younger healthcare providers (less than 30 years) were also discovered among healthcare providers aged 30–40 (5%), and 41–50 (45%). Mild depression was found in more than 20 % of all age groups included in the study.

Findings were not significant based on years of experience. Mild depression was the highest (25.4%) among healthcare providers with 11–20 years of experience. Moderately severe depression was also the highest (5.5%) among the same group. Severe depression (5.1%) was mostly noted among healthcare providers with less than 5 years of experience.

For occupation, residents scored the highest in mild, moderate, and severe depression at 33.3%, 13.9 %, and 5.6% respectively. Moderately severe depression was mostly reported among Pharmacists and nurses at 4.9% and 4.8%, respectively.

Among different specialties, psychiatry scored the highest in both moderately severe (10%) and severe (5%) depression categories. For moderate depression, pediatricians exhibited the highest rate (13.7%) compared to other specialties. Internal medicine and Obstetrics and gynecology were subspecialties that most reported mild depression and with similar rates (26.5% and 26.5%). P value was not significant.

Among various practice health care providers working in inpatient based care, specialists scored highest in the mild, moderate, moderately severe, and severe depression categories (12%, 4.7%, 1.8 % and 1.1%, respectively). P value was 0.05 (See Table 3).

**Table 3:** **Depression severity scores and prevalence among healthcare providers in correlation with HCP gender, years of experience, age group, occupation, and specialty.**

|  |  |  |  |  |  |
| --- | --- | --- | --- | --- | --- |
| Variable | Mild depression (%) | Moderate depression (%) | Moderate severe depression (%) | Severe depression (%) | P value |
| **Gender** |  |  |  |  | 0.119 |
| Female HCP | 23.5 | 9.1 | 3.6 | 2.5 |  |
| Male HCP | 19.8 | 8.8 | 6.6 | 3.5 |  |
| **Years of experience** |  |  |  |  | 0.088 |
| less than 5 | 22.7 | 11.9 | 4 | 5.1 |  |
| 6-10 | 19 | 10.6 | 3.9 | 2.3 |  |
| 11-20 | 25.4 | 8.1 | 5.5 | 3 |  |
| 21-30 | 22.4 | 7.1 | 3.1 | 1.5 |  |
| more than 30 | 16.1 | 5.4 | 1.8 | 0 |  |
| **Age group** |  |  |  |  | 0.119 |
| less than 30 | 23.9 | 13.2 | 4.4 | 4.4 |  |
| 30-40 | 22.9 | 10.1 | 5 | 3.1 |  |
| 41-50 | 22.7 | 6.5 | 4.5 | 2.3 |  |
| more than 50 | 20.3 | 7.1 | 2.4 | 1.4 |  |
| **Occupation** |  |  |  |  | 0.304 |
| consultant | 22.2 | 13.3 | 4.4 | 0 |  |
| Nurse | 22.2 | 8.8 | 4.8 | 3 |  |
| pharmacist | 23 | 11.5 | 4.90 | 0 |  |
| Resident | 33.3 | 13.9 | 0 | 5.6 |  |
| Specialist | 30.2 | 6.7 | 3.4 | 2.7 |  |
| Technician | 15.6 | 8.9 | 3.9 | 2.8 |  |
| **Specialty** |  |  |  |  | 0.351 |
| Family medicine | 23.1 | 9 | 2.6 | 1.3 |  |
| Internal medicine | 26.5 | 13.3 | 6.1 | 1 |  |
| Obstetrics and gynecology | 26.5 | 12.7 | 1 | 2.9 |  |
| Pediatric | 21.6 | **13.7** | 3.6 | 2.9 |  |
| Psychiatry | 20 | 5 | **10** | **5** |  |
| Surgery | **28.3** | 5 | 4.2 | 3.3 |  |
| Others | 20.6 | 7.7 | 4.8 | 2.9 |  |
| Overall | 11.9 | 4.7 | 2.3 | 1.4 |  |
| **Practice sitting** |  |  |  |  | 0.05 |
| Emergency and ICU care | 3.4 | 1.7 | 1.2 | 0.6 |  |
| In-patient hospital | 12 | 4.7 | 1.8 | 1.1 |  |
| Other | 2.9 | 1.3 | 0.9 | 0.6 |  |
| Primary health care | 4.3 | 1.4 | 0.4 | 0.4 |  |
| Depression scoring system included 9 items. Each item has four response. Each response had a certain score as the following: not at all=0, several days=1, more than half a day=2, almost every day=3.<br>Depression items total score ranged from 0-27. Total score of 0-4 indicate minimal depression. Total score of 5-9 indicate mild depression. Total score of 10-14 indicate moderate depression. Total score of 15-19 indicate moderately severe depression. Total score of 20-27 indicate severe depression |  |  |  |  |  |

Linear regression for anxiety score showed it negatively associated with age and working in primary health care P value of (0.002), (0.005) respectively and B= (−0.89), (−0.08) respectively. Linear regression for depression score showed it negatively associated with age, p = .000 and B= −0.125. For other sociodemographic variables, no significant associations were found. (Table 4)

**Table 4:** Linear regression analysis for factors associated with depression and anxiety.

|  |  |  |  |  |
| --- | --- | --- | --- | --- |
|  |  | Beta | t | Sig. |
| Depression Score | Age | -0.125 | -4.433 | .000 |
| Anxiety score | Age | -0.89 | -3.148 | .002 |
|  | primary health care | -0.08 | -2.826 | .005 |

Comparing the anxiety mean score among different resident specialties showed the obstetrics and gynecology residents exhibited the highest mean score for anxiety (20) followed by internal medicine 9.8. While family medicine and pediatric had lower score (3.7).

Similar findings for mean depression score were found to be highest among obstetrics and gynecology residents (21), followed by internal medicine (8.1). While family medicine, surgery and pediatric were (3.4, 4.5, 3.5) respectively.

## Discussion

The prevalence of depression and anxiety among healthcare professionals in SEHA is high during the time of the COVID-19 pandemic (38.3 %, 51.5 %) respectively. Results of this study are supported by systemic review and met analysis, which relieved a pooled prevalence of anxiety is 23.2%, and a depression prevalence rate of 22.8% although this study revealed higher anxiety prevalence.^6^ Similar presence of sever anxiety was detected in another study conduct in UAE during pandemic, however milder and moderated cases were more in this study. this can be attributed to different scales used.^4^ Que J, et al., demonstrated that prevalence of anxiety and other psychological problems were higher among frontline healthcare workers, compared with healthcare worker who did not participate in frontline work.^7^ This was further demonstrated in the present study, with severe anxiety noted among healthcare providers within the internal medicine specialty. Additionally, health care workers in inpatient settings revealed more depression among medical professionals than other settings.

Males registered more moderately severe and severe depression than females, though findings do not support the indication that female healthcare and nursing staff reported higher rates of anxiety and depression than other workers.^6^

Among respondents, residents experienced the highest rates of severe anxiety and severe depression, while obstetricians and gynecologists achieved the highest mean score for depression and anxiety. Psychological support and training are necessary for residents, to ensure their wellbeing, and intervention is required for obstetrics and gynecology residents.

Initiative-taking measures and mental health support are needed to ensure psychological wellbeing among healthcare providers.^8^Although healthcare providers in Abu Dhabi were supported by numerous measures, including the provision of psychological counseling, mental health support helplines, psychological motivational online webinar frontline specialties should receive special attention regarding this matter. Coping with stress and establishing strategies for resilience, are imperative for health care providers. Abu Dhabi was rated among the best performing cities in the world with regards to preparedness and supportive measures among them, the healthcare system. Nevertheless, the mental health of the healthcare workers seems an area worth investing at with the high depression and anxiety found. In interventional studies conducted in the United Kingdom (UK), the provision of an e-package for healthcare workers that includes evidence-based guidance, support and signposting related to psychological wellbeing was mandated for healthcare employees. ^9^ This study can be the first step in preparing action plan to support the healthcare workers mental health, but designing any intervention will need a qualitative study to ensure successful outcome. Vital component of the disaster management of infectious pandemics is to address the psychological effects in health care provider.^10^

## Conclusion

HCP experienced a high prevalence of anxiety and depression during the COVID-19 pandemic, suggesting that mental health intervention and support are necessary for HCP to ensure the psychological wellbeing of health care providers.

## Data Availability

Data available on request, all important information mentioned in the manuscript

## Limitations of the study

The participants in this study primarily originated from Abu Dhabi city and represent Abu Dhabi healthcare population. This, therefore, limited the generalizability of the study findings, although the respondents originated from both, public and private sectors. Social desirability bias may have occurred, because the questionnaire was self-administered.

However, the anonymity of the questionnaires was maintained. Despite limitations, the study provides a reasonable source of information. Follow up studies to assess the progression of the psychological impact of COVID −19 pandemic is needed. No medically recorded review was accomplished for the health care providers, and social factors and support were not measured in this study. The timing of the study is at the earliest stages of the pandemic and this may have differed later.

## Abbreviations

COVID-19: Coronavirus Disease-2019
WHO: World Health Organization
KAPs: Knowledge, Attitudes and Practices
HCW: Healthcare Worker
HCPs: Healthcare providers
HCP: Healthcare Professionals
PHQ-9: Patient Health Questionnaire-9
GAD 7: Generalized Anxiety Disorder −7

## Availability of data and material

All data are fully available without restrictions. All relevant data are within the paper and supplementary files.

## Authors’ contributions

**AKA, LMA, HMA, RAA :** conception and design, acquisition of data.

**AKA**, **AAA, LBA :** Analysis and interpretation of data. Drafting the article or substantively revising it critically for important intellectual content.

All authors read and approved the final manuscript.

## Acknowledgements

Not applicable

## Ethics approval and Consent to participate

The study was approved by the Ambulatory Health care services human ethics committee and the Abu Dhabi health care service central human Ethics Committee. Consent was obtained from participants. All participants were fully informed of every aspect of the study, and participation was completely voluntary. Information provided by the participants was recorded anonymously, and has been kept confidential.

